# Rapid Species-Specific Molecular Diagnosis of Amoebic Liver Abscess by Isothermal RNA Amplification: A Case Report

**DOI:** 10.64898/2026.11.24.26368577

**Authors:** Hércules Pereira Neves, Antônio Rogério Dias Guimarães, João Francisco Leão dos Santos, Milena Martins Magnus, Vinicius Ribeiro Silva, Raquel Bandeira da Silva, Glauco Sobreira Messias, Wagner Eduardo Ferreira

## Abstract

**Background:** Amoebic liver abscess, caused by *Entamoeba histolytica*, is a potential life threatening extra intestinal infection that is prevalent in tropical and subtropical regions. *Entamoeba histolytica* is morphologically indistinguishable from the non-pathogenic *Entamoeba dispar*, yet species-level confirmation is critical before initiating metronidazole. This is particularly important in elderly patients, for whom the drug carries a higher risk of QT prolongation, peripheral neuropathy, and hepatotoxicity. In Brazil, where *E. dispar* predominates over *E. histolytica* at an estimated 10:1 ratio, diagnostic misidentification constitutes a genuine hazard.

**Methodology and Principal Findings:** We report a patient with amoebic liver abscess with secondary hepatic biochemical derangement requiring urgent molecular species confirmation. A Nucleic Acid Sequence-Based Amplification (NASBA) assay was designed targeting a species-specific region of the *E. histolytica* 40S small subunit ribosomal RNA (GenBank MW026793.1). Primer design using NCBI Primer-BLAST yielded a thermodynamically balanced primer pair, with melting temperatures of 53.25°C and 53.81°C and a resulting amplicon with 121 nucleotides, which is suitable for standard NASBA at 41°C. Applied to an aspirated abscess sample, the assay returned a positive result confirming *E. histolytica* infection and enabling continuation of antiamoebic therapy with favourable clinical outcome.

**Conclusions and Significance:** NASBA targeting *E. histolytica* small subunit ribosomal RNA provides species-specific diagnosis through two orthogonal mechanisms: sequence-level discrimination from *E. dispar* at the primer binding sites, and the intrinsic viability bias or RNA amplification, which favours signal from metabolically active trophozoites over residual nucleic acid. The episomal architecture of *E. histolytica* ribosomal DNA, which has approximately 200 extrachromosomal gene copies per trophozoite (each actively transcribed), confers inherent sensitivity amplification. Operating isothermally at 41°C without a thermocycler or any additional laboratory infrastructure, this approach addresses a genuine diagnostic gap in neglected tropical disease management and is suited to point-of-care deployment in resource-limited endemic settings.

**Author Summary:** We work in Brazil, where two closely related parasites, *Entamoeba histolytica* and *Entamoeba dispar*, create a diagnostic problem with real consequences. They look identical under the microscope, but only *E. histolytica* causes disease, including dangerous liver abscesses. The standard treatment is based on metronidazole, which can harm elderly patients if used unnecessarily, yet withholding it in a true infection risks life threatening progression. Telling the two species apart rapidly matters; however, molecular tests capable of doing so require equipment and laboratory infrastructure unavailable in most tropical settings.

We designed a test that explores a feature unique to *E. histolytica*: its instructions for building ribosomes are stored on hundreds of small circular DNA molecules outside the chromosomes of the parasite, all of which continuously active. This makes ribosomal RNA an extraordinarily abundant target. Our test amplifies this RNA at a single fixed temperature using only a basic heating block. It was able to confirm *E. histolytica* in a patient with severe liver abscess and justified the continuation of treatment. We believe this approach could serve as a point-of-care tool in tropical regions where amoebiasis is common and diagnostic resources are scarce.

## 1. Introduction

### 1.1. Epidemiology and clinical burden of amoebiasis

Amoebiasis ranks among the most significant parasitic diseases affecting human populations globally. *Entamoeba histolytica* is estimated to infect 50 million people annually, causing 40,000 to 100,000 deaths each year, thus placing it second only to *Plasmodium falciparum* malaria as a protozoan killer [1,2]. The disease burden is disproportionately borne by populations in sub-Saharan Africa, South and Southeast Asia, and Latin America, where inadequate water sanitation, overcrowding, and limited healthcare access sustain transmission through the faecal-oral route via cyst-contaminated food and water [3].

The clinical spectrum of amoebiasis is broad. Approximately 90% of infected individuals remain asymptomatic intestinal carriers, while the remaining 10% develop symptomatic disease ranging from amoebic colitis (characterised by bloody diarrhoea and abdominal cramping) to invasive extra-intestinal disease [4]. Amoebic liver abscess is the most frequent and clinically serious extra-intestinal manifestation, accounting for the majority of amoebiasis-related deaths. When left untreated, amoebic liver abscess carries a mortality rate in excess of 5%, rising sharply with rupture into the pleural or pericardial space [5]. Men aged 18 to 50 years are disproportionately affected, although the outcomes are substantially worse in immunocompromised patients and the elderly, in whom impaired cellular immunity removes a critical barrier to trophozoite invasion [6].

In Brazil, amoebiasis remains a public health concern, particularly in regions such as the Amazon basin and settlements with inadequate sanitation. National prevalence data are complicated by the historical failure in distinguishing *E. histolytica* from *E. dispar* in surveillance reports [7]. Contemporary molecular surveys suggest that true *E. histolytica* infection accounts for approximately 10% of all *Entamoeba*-positive cases in most Brazilian cohorts [8]. The epidemiological reality underpins the diagnostic challenge at the centre of the present case: the *a priori* probability that any given *Entamoeba*-positive patient harbours the non-pathogenic species is approximately 90%, which makes empirical antiamoebic therapy a statistically unjustifiable default in a clinical setting where species discrimination is feasible.

### 1.2. The diagnostic dilemma: *E. histolytica* vs. *E. dispar*

The similarity between *Entamoeba histolytica* and *Entamoeba dispar* is staggering. Their cysts and trophozoites are indistinguishable under the conventional light microscope: their size, nuclear architecture, and cytoplasmic appearance. This fact was recognised by Brumpt as early as 1925 but not formally acted upon until their re-description by Diamond and Clark in 1993 [7,9]. For nearly seventy years the two organisms were conflated under a single species name, leading to the systematic overestimation of invasive amoebiasis prevalence and unnecessary treatment of millions of asymptomatic *E. dispar* carriers [3,4].

The clinical consequences of this distinction are asymmetric and serious. *Entamoeba dispar* is genuinely non-pathogenic: it colonises the intestinal lumen without invading tissue, does not cause colitis or hepatic disease, and requires no treatment [3,4]. *Entamoeba histolytica*, by contrast, is an obligate pathogen in those who develop invasive disease, capable of mucosal invasion, haematogenous spread, and hepatic abscess formation with the potential for rupture and death [2,4]. A false positive identification of *E. histolytica* therefore exposes the patient to unnecessary metronidazole without any parasitological benefit.

The risks of unnecessary metronidazole are not trivial, particularly in the elderly. The drug is known to prolong the cardiac QT interval, interacts with alcohol in a reaction similar to disulfiram, and causes dose-dependent peripheral neuropathy with prolonged courses [10]. Hepatotoxicity is an additional concern in patients already experiencing hepatic compromise. In elderly patients with concurrent liver disease, these risks are amplified by age-related pharmacokinetic changes, polypharmacy, and reduced physiological reserve.

Conversely, withholding drug treatment in a patient with confirmed *E. histolytica* amoebic liver abscess carries its own mortality risk. Untreated amoebic liver abscess progresses to rupture in a substantial number of cases, with peritoneal, pleuropulmonary, or pericardial extension carrying mortality rates of 20 to 40% [2,5]. The diagnostic dilemma is therefore not a matter of academic precision but of competing therapeutic risks: the harm of treating *E. dispar* versus the harm of not treating *E. histolytica*. Species-level molecular confirmation resolves this dilemma definitely and is the rational basis for the approach described in this report.

### 1.3. Limitations of conventional diagnostic approaches

The accurate diagnosis of *Entamoeba histolytica* infection in the clinical setting remains difficult; none of the conventional approaches available at most points of care provides confirmation at the species level with adequate sensitivity and specificity for amoebic liver abscess. Microscopy of stool and aspirate material is the oldest and most widely used method in endemic settings but is compromised by two basic limitations. First, it cannot distinguish *E. histolytica* from *E. dispar* or *E. moshkovskii* (a third morphologically identical species of uncertain pathogenicity that has emerged as an increasingly recognised human commensal [3]. Second, its sensitivity for diagnosing amoebic liver abscess is poor, given that fewer than half of the amoebic liver abscess patients have detectable intestinal *Entamoeba* at the time of presentation, meaning a negative stool examination carries no diagnostic weight in this context [3,4]. The sensitivity of direct microscopy for amoebic liver abscess diagnosis is estimated at 30 to 40% at best and high inter-observer variability in morphological interpretation further limits its reliability [11].

Serology is a useful adjunct for amoebic liver abscess diagnosis and can achieve sensitivities of 70 to 90% in established disease [3,12]. However, it has critical limitations in acute clinical decisions. Antibody titres take 5 to 7 days to rise to detectable levels after the onset of invasive infection; this makes serology unreliable in the first week of disease, which happens to be the window during which rapid treatment decisions are most consequential [12]. Furthermore, seropositivity persists for years after the clearance of infection, therefore a positive result cannot distinguish current active amoebic liver abscess from an old exposure, particularly in individuals from endemic regions [3]. Serology is also unable to differentiate a new infection from recurrence.

Antigen detection enzyme immunoassays detect the species-specific galactose-inhibitable adherence lectin in stool and serum and offer genuine species discrimination with sensitivities of over 80% for intestinal disease [3,13]. However, their performance in liver abscess aspirate is substantially lower, reportedly as poor as 33% following even brief metronidazole pre-treatment [13]. This is a common scenario in patients who have already received empirical therapy before specialist assessment.

Real-time PCR is the current molecular gold standard. Its sensitivity and specificity exceed 95% with simultaneous species discrimination [3,14]. However, it requires complex and expensive laboratory infrastructure that remains unavailable at most points of care in regions where amoebiasis is endemic. The situation is further complicated by long turnaround times, particularly when clinical decisions are urgent.

### 1.4. NASBA as a molecular diagnostic platform

Isothermal amplification constitutes a powerful tool for molecular diagnosis that brings together simplicity, robustness, quality, and speed, without losing the well-known attributes of PCR [15]. It is particularly suited for field use or wherever laboratory infrastructure is poor or non-existent. There are more than 15 different isothermal amplification techniques, thus enabling the choice of the most suitable options in each context. Nucleic Acid Sequence-Based Amplification (NASBA) is an isothermal amplification technology developed by Compton in 1991 for the amplification of RNA [16]. It operates at a single fixed temperature of 41°C and is driven by three enzymes acting simultaneously: avian myeloblastosis virus reverse transcriptase (AMV-RT), ribonuclease H (RNase H), and T7 RNA polymerase. In the initiation phase, the antisense primer (which carries the T7 RNA polymerase promoter sequence at is 5’ end) anneals to the target RNA and is extended by AMV-RT to generate a cDNA strand. Next, RNase H selectively degrades the RNA component of the RNA:cDNA hybrid. The sense primer then anneals to the single-stranded cDNA and AMV-RT completes the double-stranded DNA intermediate. A cyclic phase follows suit, where T7 RNA polymerase binds the promoter region and transcribes up to 1,000 antisense RNA copies per double-stranded DNA template; each of them re-enters the cycle as a new template. The result is amplification of more than 10^12^ copies of the target sequence within 90 to 120 minutes without any temperature cycling [17,18].

Two properties of NASBA are particularly relevant in the present context. First, it is selective for RNA. Given that the reaction works isothermally without the denaturation step that characterises PCR, double stranded genomic DNA remains in its native conformation and is inaccessible to the antisense primer. NASBA therefore amplifies RNA directly and exclusively, without generating false-positive signals from residual genomic DNA contamination. This is a particularly important advantage in the analysis of complex clinical matrices such as liver abscess aspirate or serum, where host DNA is abundant [18]. Secondly, its isothermal character requires only a simple heat block or water bath. This positions NASBA as a technically feasible platform for molecular diagnosis in primary care facilities and hospitals lacking sophisticated laboratory infrastructure.

NASBA has been successfully applied to the molecular diagnosis of a range of infectious diseases, including HIV-1 viral load quantification, respiratory virus detection, and the diagnosis of parasitic infections including *Trypanosoma brucei* [18,19]. Its application to *Entamoeba histolytica*, and specifically to the high copy number episomal SSU ribosomal RNA target described herein, constitutes a logical extension of the platform to a clinically compelling neglected tropical disease for which rapid point-of-care diagnosis remains an unmet need.

### 1.5. Study objectives

We report the case of a patient presenting with amoebic liver abscess with secondary cholestatic hepatic biochemical derangement that required rapid species-level molecular confirmation to support continuation of antiamoebic therapy. We describe the design and application of a NASBA assay targeting a species-specific region of the *Entamoeba histolytica* 40S small subunit ribosomal RNA. The molecular rationale underpinning the choice of this target is also presented. This includes the episomal architecture of *E. histolytica* ribosomal DNA that sustains exceptionally high RNA abundance per trophozoite and the dual specificity mechanisms that underpin the assay: sequence discrimination at the primer binding sites (which exploits the nucleotide divergence between *E. histolytica* and *E. dispar* SSU rRNA) and the mechanistic RNA selectivity inherent to NASBA amplification (which excludes false-positive signal from any genomic DNA).

## 2. Case Presentation

A male patient aged within the 60-70 range with a background of systemic arterial hypertension, managed with amlodipine, losartan, and atenolol, presented with a three-day history of acute right hypochondrium pain of pleuritic character, associated with a dry cough. He reported worsening of the pain on deep inspiration. There was no history of tobacco use, drug allergies or gastrointestinal symptoms at the time of initial presentation.

The patient sought care at an urgent care unit, where the initial working diagnosis was community-acquired pneumonia. Empirical treatment was commenced with amoxicillin, subsequently escalated to amoxicillin-clavulanate combined with azithromycin, without clinical improvement. Laboratory investigations at the time demonstrated leucocytosis (18,100/mm^3^), markedly elevated C-reactive protein (195,9 mg/L), and mildly elevated hepatic enzymes (AST 73 U/L, ALT 96 U/L, GGT 368 U/L), with preserved renal function. Chest radiography showed consolidation with right basal pleural effusion.

The patient was transferred to the general hospital for diagnostic investigation and management. On admission, he was haemodynamically stable and afebrile. Physical examination revealed reduced vesicular breath sounds at the right lung base; the abdomen was non-tender with no signs of peritoneal irritation.

Computer tomography of the thorax and abdomen (Figure 1) demonstrated a small right pleural effusion with compressive atelectasis and an intrahepatic collection in the right lobe measuring approximately 8.6 × 4.1 × 4.4 cm (estimated volume 80.7 cm^3^), consistent with a hepatic abscess, with associated stranding of the adjacent fat and thickening of the peritoneal reflection. Irregular thickening of the fundal wall of the gallbladder was also noted. Abdominal ultrasonography (Figure 2) confirmed a liver of normal volume, shape, and echotexture with moderate diffuse increase in echogenicity and identified an elongated cystic image with irregular contours, well-defined margins, and internal echoes without Doppler vascularisation, measuring approximately 10.0 × 3.5 × 4.5 cm (estimated volume 83.5 cm^3^).

**Figure 1.**
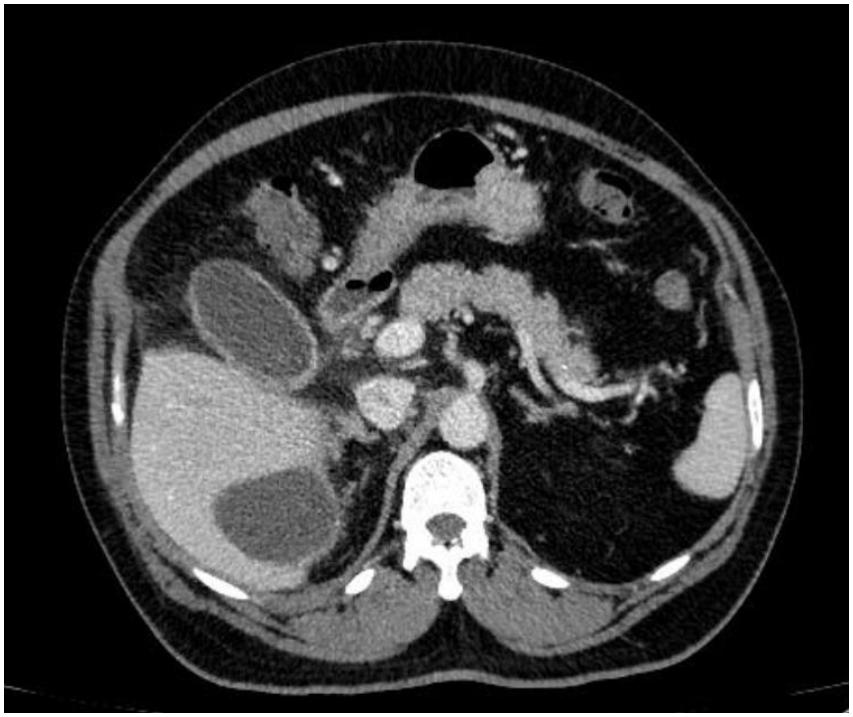
Abdominal computer tomography demonstrating an intrahepatic cystic lesion measuring approximately 8.6 × 4.1 × 4.4 cm (volume 80.7 cm^3^).

**Figure 2.**
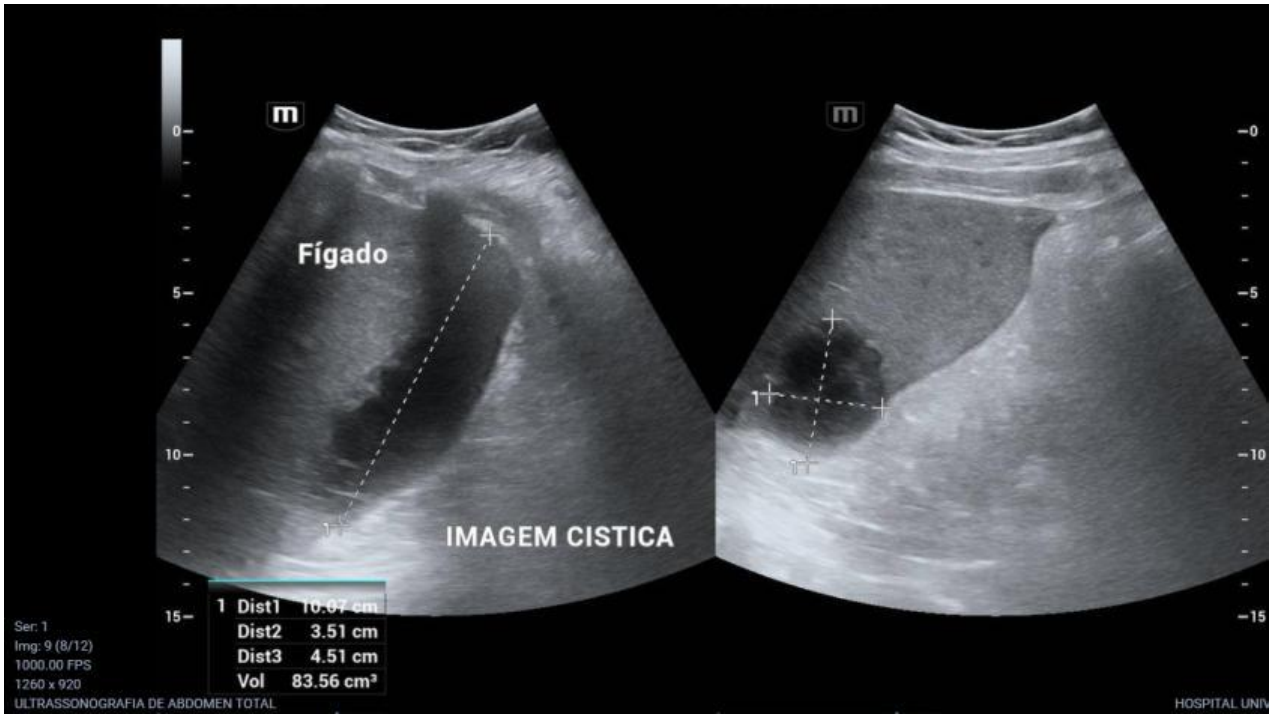
Abdominal ultrasonography demonstrating an intrahepatic cystic lesion measuring approximately 10.0 × 3.5 × 4.5 cm (volume 83.5 cm^3^).

The working diagnosis was hepatic abscess of undetermined aetiology: amoebic versus pyogenic. Empirical treatment was initiated with intravenous ceftriaxone 1 g every 12 hours (planned for four weeks), metronidazole 500 mg every 8 hours for seven days, and tinidazole 2 g once daily for five days.

The patient underwent percutaneous ultrasound-guided aspiration of the hepatic lesion, which yielded turbid material with particulate matter; continuous drainage was not required. Microbiological analysis of the aspirate showed no bacteria on Gram staining and a negative general culture. Microscopy of the aspirate did not identify *Entamoeba* trophozoites. Stool parasitological examination was negative. Serological investigations for HIV, viral hepatitis, and syphilis were all negative.

In the absence of a microbiological or morphological diagnosis and given the clinical and radiological picture strongly suggestive of amoebic liver abscess, the aspirate material was submitted for NASBA amplification targeting the *Entamoeba histolytica* 40S small subunit ribosomal RNA, as described in Section 3. The assay yielded a positive result, confirming *E. histolytica* infection and providing the species-level molecular diagnosis that conventional investigations had been unable to establish.

## 3. Molecular Methods

### 3.1. Target selection: the *E. histolytica* 40S small subunit Ribosomal RNA

The selection of the *E. histolytica* 40S small subunit (SSU) ribosomal RNA as our NASBA target was determined by three converging properties: exceptionally high copy number, active transcription in viable trophozoites, and the presence of species-specific sequence signatures that discriminate *E. histolytica* from *E. dispar* with the primer binding region.

Unlike virtually all other eukaryotes, *E. histolytica* does not encode its ribosomal RNA genes on chromosomal DNA. Instead, the complete ribosomal DNA unit (which comprises the SSU rRNA, 5.8S rRNA, and large subunit (60S) rRNA genes along with their intervening and flanking sequences) is organised on extrachromosomal palindromic circular DNA episomes of approximately 25 kilobases [20]. This arrangement bears striking similarity with bacterial plasmids: circular, extrachromosomal, autonomously replicating with their own replication origin, relatively stable copy number (which is regulated independently of chromosomal replication), and palindromic organisation. Each episome carries two inverted copies of the rDNA unit arranged symmetrically around a central AT-rich region and each trophozoite maintains approximately 200 such episomes, yielding roughly 400 ribosomal gene copies per cell [20,21]. All of these copies are actively transcribed, generating a steady abundance of 40S rRNA molecules that is orders of magnitude greater than would be produced from a single chromosomal locus. This episomal architecture functions as an endogenous amplification system, where the target molecule is present at high concentration before the NASBA reaction even begins.

The choice of targeting RNA rather than the rDNA episome is deliberate and mechanistically relevant. NASBA operates isothermally at 41°C without a denaturation step. This means that double-stranded DNA (including the episomal rDNA) remains in its native conformation and is inaccessible to the antisense primer. The assay therefore amplifies the mature 40S rRNA transcript exclusively, generating no signal from the episomal DNA contamination regardless of how much host or parasite genome material might be present in the sample matrix [17,18]. In the context of a liver abscess aspirate, where host DNA is abundant and the sample matrix is complex, such selectivity for RNA constitutes an important analytical advantage over PCR-based approaches.

A second layer of specificity is conferred at the sequence level. The 40S SSU rRNA of *E. histolytica* diverges from that of *E. dispar* by approximately 1.7% at the nucleotide level [22] and the primer binding sites selected for this assay span a signature region that exploits this divergence. The antisense primer NP1 anneals exclusively to *E. histolytica* SSU rRNA sequence and does not bind to *E. dispar, E. moshkovskii*, or human ribosomal RNA under standard NASBA conditions. Specificity for *E. histolytica* over *E. dispar* is therefore achieved through two independent mechanisms: sequence discrimination at the primer binding site and mechanistic discrimination through RNA selective amplification. Neither of them is available to DNA-targeted assays, which must rely on sequence differences alone.

The reference sequence used for primer design was GenBank accession number MW026793.1 (*Entamoeba histolytica* isolate 1110, 40S SSU rRNA gene, partial sequence).

### 3.2. Species discrimination at the sequence level

The species discrimination capacity of the assay rests on the well-characterised sequence divergence between *E. histolytica* and *E. dispar* 40S SSU rRNA genes within the target region. Although the two species are morphologically indistinguishable and share a recent common evolutionary ancestor, their SSU rRNA sequences diverge by approximately 1.7% at the nucleotide level across the gene as a whole [22], with the divergence concentrated in specific hypervariable regions interspersed between conserved structural helices. Such organisation is precisely what allows a single primer pair to anchor to a shared structural context while exploiting a locally divergent sequence for discrimination.

The genus *Entamoeba* presents a diagnostic challenge that extends beyond the *E. histolytica* and *E. dispar* pair. *E. moshkovskii*, a third morphologically identical species, is now recognised as a genuine human commensal and has been identified in stool specimens from patients with gastrointestinal symptoms across multiple countries [3]. Its SSU rRNA sequence diverges sufficiently from *E. histolytica* to permit species-specific primer design; published primer sets targeting the *E. histolytica* SSU rRNA have been shown to be non-reactive with *E. moshkovskii* [22]. Additional non-pathogenic species found in clinical samples include *E. hartmanni, E. coli, E. poecki*, and *Endolimax nana*, all of which differing from *E. histolytica* at the molecular level within this region, therefore excluded by species-specific primers.

The BLAST analysis described in Section 3.3 confirmed empirically that the NP1 and NP2 gene-specific sequences do not align to any non-*histolytica Entamoeba* species in GenBank, thus providing *in silico* support for the sequence-level specificity argued here. The clinical importance of this molecular discrimination deserves special emphasis: although *E. dispar* does not disseminate to the liver and is not expected to be present as an invasive pathogen in abscess material, the aspiration procedure itself carries a risk of contamination with intestinal or peritoneal contents harbouring commensal *E. dispar* trophozoites or cysts. In this scenario, sequence level discrimination at the primer binding site constitutes the essential safeguard against a false-positive result. The two layers of specificity described in this report are therefore not redundant but complementary: biological exclusion based on pathogenic behaviour provides the epidemiological rationale for testing, while sequence-level primer discrimination provides the analytical guarantee of a correct result even in the presence of sample contamination.

### 3.3. Primer design

Following thorough analysis of the 40S SSU rRNA of *E. histolytica* based mostly on GenBank accession number MW026793.1, primer design was done using NCBI’s Primer BLAST. Design parameters were set to favour NASBA compatibility: amplicon size constrained to 100 to 200 nucleotides, primer melting temperatures in the range of 50°C to 65°C under standard PCR buffer conditions, and absence of dimer or self-dimer formation. The final primer pair selected was:

NP1: **5’-**AATTCTAATACGACTCACTATAGGG**-ATACAAACTGGATCGTCTCA-3’**

NP2: **5’-AGACCAAGTAGGATGAAACT-3’**

Where the 25-nucleotide prefix in NP1 is the T7 tag for binding with the T7 RNA polymerase. The expected amplicon size, measured between the outer edges of the primer binding sites, is 121 nucleotides.

Thermodynamic parameters calculated by Primer BLAST under PCR buffer conditions (incorporating magnesium correction) were: NP2 melting temperature of 53.25°C, NP1 melting temperature of 53.81°C, and a resulting Δ*T*^m^ of 0.56°C. Such near-identical thermal stability of the two gene-specific portions ensures balanced primer annealing efficiency during the cycling phase of NASBA amplification, where both primers compete for their respective templates simultaneously at 41°C. Each primer carries a margin of approximately 12 to 13°C above the reaction temperature, which is adequate for stable and productive annealing throughout the amplification.

Following primer selection, the gene-specific sequences of both NP1 and NP2 were submitted independently to NCBI BLAST (blastn, core nucleotide database) with the search restricted to the genus *Entamoeba*, to evaluate specificity across related species. Both primers returned significant alignments exclusively to *E. histolytica* accessions, with no matches to *E. dispar, E. moshkovskii, E. hartmanni, E. coli*, or *Endolimax nana* sequences. These results confirm that both primer binding sites reside within the species-specific signature region of the *E. histolytica* 40S SSU rRNA and that cross-reactivity with morphologically similar non-pathogenic *Entamoeba* species will not occur under standard conditions.

### 3.4. NASBA reaction and detection

NASBA reactions were performed in a total volume of 20µL using an in-house assembled enzyme preparation and reaction buffer. The reaction buffer contained 40 mM Tris-HCl (pH 8.5), 12 mM MgCl2, 70 mM KCl, 5 mM dithiothreitol, 1 mM each dNTP, 2 mM each rNTP, 0.2 µM each primer, and 15% v/v dimethylsulphoxide. Nucleic acid extract from the clinical sample was added to a final volume of 5 µL per reaction. The enzyme mixture comprising avian myeloblastosis virus reverse transcriptase (AMV-RT, 6.4 units), ribonuclease H (RNase H, 0.08 units), and T7 RNA polymerase (32 units) was prepared separately and added to the reaction only after the denaturation-annealing step described below.

Prior to enzyme addition, the assembled reaction mix containing the primers, buffer, and nucleic acid extract was subjected to a denaturation step at 65°C for 5 minutes. This step serves to unfold secondary structures in the RNA target molecule that would otherwise impede primer access and is distinct in purpose from the DNA denaturation step used in PCR: at 65°C double-stranded DNA remains substantially intact and no DNA strand separation occurs. The reaction was then cooled to 41°C for 5 minutes to allow primer annealing to the single-stranded RNA template before the in-house enzyme mixture was added. Following enzyme addition, the reactions were incubated at 41°C for 90 minutes in a heated dry block.

An exogenous internal amplification control was included in every reaction to monitor for inhibition and to confirm the integrity of the in-house enzyme mixture. The control target was a synthetic RNA segment derived from the rbcL gene (encoding the large subunit of ribulose-1,5-biphosphate carboxylase/oxygenase, RuBisCO) of the Japanese kelp *Saccharina japonica* (GenBank OR228416.1). This gene was chosen specifically because the RuBisCO large subunit is encoded exclusively in the plastid genomes of photosynthetic organisms and is entirely absent from the human genome, from all animal genomes, and from all known bacterial, viral, and parasitic pathogens. This makes cross-reactivity with any component of a clinical sample from an infected patient a biological impossibility. The control RNA was introduced at a defined input copy number calibrated to produce a clearly visible band at a size distinct from the 121 nt *E. histolytica* amplicon. Failure of the control band in the absence of a positive signal was interpreted as evidence of reaction inhibition, invalidating the result and requiring repeat analysis with fresh extract.

Amplification products were resolved in this study by electrophoresis on a 2.5% agarose gel in 1× TBE buffer, stained with SaferDye, and visualised under UV transilumination. A 100 bp DNA ladder was included in each run as an approximate size reference for the RNA amplicons. Interpretation criteria were defined prospectively: a band at 121 nt in the presence of a visible control band constituted a positive result for *E. histolytica*; absence of the 121 nt band with a present control band constituted a negative result and absence of both bands required repeat testing.

It should be noted that gel electrophoresis was selected as the readout method for this proof-of-concept application, but NASBA amplicons are fully compatible with a range of alternative detection formats that may be more appropriate for point-of-care deployment. Lateral flow assay (LFA) strips can provide a visual result without instrumentation within minutes of amplification completion [19]. Molecular beacon probes, added directly to the NASBA reaction, enable real-time fluorescence monitoring without post-amplification tube opening, eliminating the carryover contamination risk that is inherent in gel-based workflows [23]. The choice of detection method does not alter the primer sequences, reaction chemistry, or analytical specificity of the assay; any such approaches could be substituted in future implementations targeting resource-limited settings where the isothermal character of NASBA and the infrastructure independence of LFA readout would together constitute a genuinely deployable point-of-care test.

### 3.5. Study period and ethics statement

This is a single-patient case report and no recruitment was performed. The patient was identified during routine clinical care. The study period corresponds to the patient’s hospital stay, from admission to discharge in March 2026. Ultrasound-guided aspiration of the hepatic lesion and NASBA testing of the aspirate were both performed within that period. Clinical and laboratory data were collected from the medical records at that time.

The study was approved by the Research Ethics Committee of Faculdade Ciências Médicas de Minas Gerais through Plataforma Brasil (CAAE 01932226.2.0000.5134; approval no. 8.793.352). Written informed consent for the publication of this case report, including clinical data and images, was obtained from the patient.

## 4. Results

### 4.1. NASBA assay result

Nucleic acid extraction from the liver abscess aspirate was performed using the XMagic™ fast extraction protocol. The NASBA reaction followed the preparation and protocol described in Section 3.4. Following amplification, the reactions were terminated with a stop solution for 5 minutes. Amplification products were resolved by electrophoresis on a 2.5% denaturing agarose gel prepared in MOPS buffer. Electrophoresis was carried out at 70 V and 56 mA for 60 minutes, with the gel tank kept on ice. Bands were visualised under UV transilumination. A 100 bp DNA ladder was used as a molecular size marker; this was an approximate assessment given the RNA nature of the amplification products.

The NASBA assay produced a distinct amplification product corresponding to the *Entamoeba histolytica* target. Based on its electrophoretic migration relative to the molecular size marker, the observed band was consistent with an amplicon of approximately 120 nucleotides, closely matching the predicted size. The internal amplification control generated a separate and clearly distinguishable band, supporting the absence of significant reaction inhibition and confirming preservation of enzymatic activity throughout the assay (Figure 3). The simultaneous presence of both the target-specific band and the internal control band was interpreted as a positive result for *E. histolytica*. Moreover, the absence of nonspecific bands or evident primer-dimer artefacts supports the analytical specificity of the assay under the reaction conditions.

**Figure 3.**
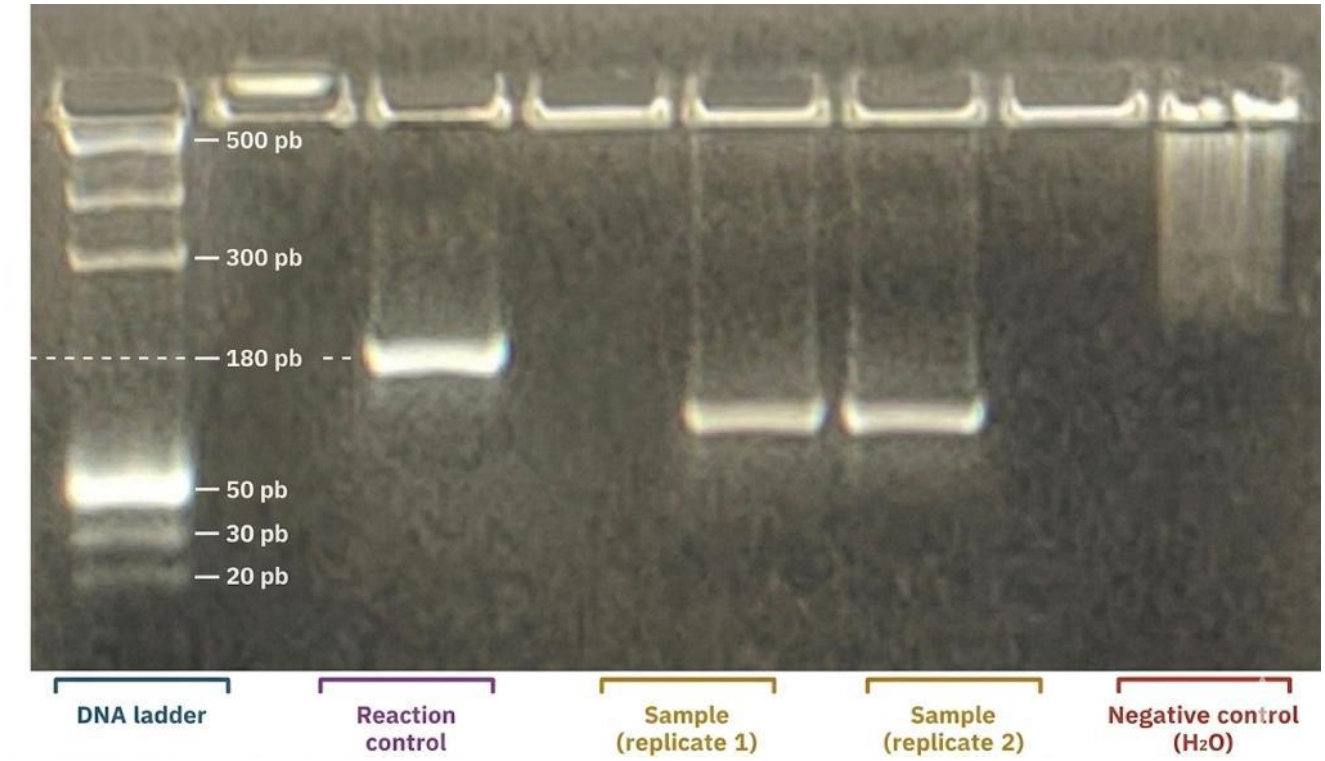
Electrophoresis gel showing the NASBA amplification results.

### 4.2. Clinical outcome

The patient had a favourable clinical course. C-reactive protein fell from 132.8 mg/L on admission to 10.2 mg/L at discharge; leucocytosis resolved; cholestasis markers and hepatic enzyme levels improved progressively. The right pleural effusion remained small and of no haemodynamic significance and was managed conservatively as a reactive exudative effusion. The patient was discharged in good general condition, continuing ceftriaxone under a home care programme with planned outpatient clinical and laboratory follow up.

## 5. Discussion

### 5.1. The clinical case for molecular species confirmation in ALA

Amoebic liver abscess presents a clinical scenario in which the consequences of diagnostic error are asymmetric and potentially fatal. Unlike intestinal amoebiasis, in which the distinction between *E. histolytica* and the non-pathogenic *E. dispar* can sometimes be deferred without immediate clinical risk, hepatic involvement demands prompt treatment decisions. Delay poses a risk of abscess progression and rupture, while unnecessary administration of metronidazole exposes the patient to drug toxicity without benefit. The argument for species-level molecular confirmation in ALA is therefore not merely academic. It is a prerequisite for rational clinical management.

The standard diagnostic approach in most endemic settings still relies on a combination of clinical and radiological criteria, serology, and microscopy. Each of these carries limitations that are amplified in the ALA context. Clinical and imaging findings are nonspecific and cannot differentiate amoebic from pyogenic or other hepatic abscesses with sufficient confidence to guide definitive treatment without microbiological support [2,5]. Serology is sensitive in established disease but unreliable within the first week of illness, when treatment decisions are most consequential [12]. Microscopy of aspirate material cannot resolve the species question, since *E. histolytica* and *E. dispar* are indistinguishable by morphology [3,7]. Antigen detection, the most promising of the conventional approaches, is acutely vulnerable to pre-analytical confounders (sensitivity drops sharply after even brief metronidazole pre-treatment) [13]. In practice, the clinical decision to treat is frequently made based on probability rather than confirmed species identification, which is acceptable in poor resource settings but constitutes a diagnostic gap that directly affects patient safety.

Molecular confirmation by NASBA targeting the *E. histolytica* 40S SSU rRNA closes this gap through a mechanism specifically suited to the ALA clinical context. Liver abscess aspirate is the ideal molecular specimen, as it is obtained directly from the pathology site, it is available as part of the therapeutic drainage, and it avoids the insensitivity of stool-based methods in patients whose intestinal *Entamoeba* burden may be low or absent [3,4]. Within this matrix, RNA-targeted isothermal amplification provides species-level discrimination at the sequence level, which is a distinction that gains particular significance because the aspiration procedure itself carries a risk of contamination with peritoneal or intestinal contents harbouring commensal *Entamoeba* species, thus making primer-level sequence discrimination the essential safeguard against false-positive identification rather than a redundant additional layer [22]. The case described in this report illustrates why that safeguard matters clinically and why the development of accessible molecular diagnostic platforms for ALA represents a significant step ahead in the management of this neglected tropical disease.

### 5.2. Why rRNA rather than rDNA: two orthogonal specificity layers

The decision to target ribosomal RNA rather than the ribosomal DNA locus from which it is transcribed is central to the design logic of this assay and warrants explicit discussion because it determines both the analytical platform that can be used and the nature of the specificity that it provides.

The episomal rDNA architecture of *E. histolytica* (which has approximately 200 extrachromosomal palindromic DNA rings, each carrying two inverted DNA copies yielding roughly 400 ribosomal gene copies per trophozoite [20,21]) means that the steady-state abundance of 40S SSU rRNA molecules in a viable cell is orders of magnitude greater than would be produced from a single copy chromosomal locus. This endogenous amplification system is analytically exploitable only if the target double-stranded DNA remains in its native helical conformation at 41°C and is physically inaccessible to the antisense primer [16,17]. A NASBA assay directed at rRNA therefore amplifies from a starting template that is both exceptionally abundant and exclusively single-stranded, combining high sensitivity with intrinsic resistance to a false-positive signal from any source of double-stranded DNA (such as episomal DNA rings themselves, residual host genomic DNA, or environmental DNA contamination).

This RNA selectivity constitutes the first specificity layer and operates mechanistically, independently of any sequence consideration. Its value in the context of a liver abscess aspirate is highly significant: the sample matrix contains an abundance of host hepatic DNA and any DNA-targeted amplification method must actively exclude this background through primer design alone. The NASBA mechanism provides this exclusion automatically and unconditionally, without placing any additional burden on primer specificity.

The second specificity layer operates at the sequence level and is independent of the first. NP1 and NP2 were selected from the hypervariable region of the 40S SSU rRNA gene where *E. histolytica* diverges from *E. dispar* by approximately 1.7% at the nucleotide level. BLAST analysis against the complete *Entamoeba* genus confirmed that neither primer sequence aligns to *E. dispar, E. moshkovskii, E. hartmanni, E. coli*, or *Endolimax nana* under standard conditions. Such discrimination is the essential analytical safeguard in the ALA context, given that aspirate collected by needle puncture is not a sterile specimen in the absolute sense and contamination with peritoneal and intestinal contents cannot be ruled out. Primer-level sequence discrimination therefore ensures that such contamination, if present, generates no signal.

The orthogonality of these specificity layers is what makes the combination analytically robust. Neither depends on the other, given that the RNA selectivity of NASBA would exclude DNA-based signals and the sequence-level discrimination of the primers would remain operative even if the reaction were converted to a DNA amplification format. Together, they define an assay whose specificity is guaranteed by two independent and simultaneously operating mechanisms; this is vital in a setting where the consequences of a false-positive result are both clinically significant and therapeutically reversible only at the cost of delayed treatment.

### 5.3. Episomal architecture as an sensitivity amplifier

The extraordinary sensitivity of NASBA-based detection of *E. histolytica* rests in part on a biological peculiarity of this organism that has no parallel among the medically important protozoan pathogens with which it is routinely compared: the complete absence of ribosomal RNA genes from its chromosomal DNA. In virtually all eukaryotes, rDNA is present in tandem chromosomal arrays at one or more fixed loci. In *E. histolytica*, it is absent from the chromosomes entirely, residing instead on the extrachromosomal palindromic circular episomes described in Section 3.1 [19,20]. This is not a mere curiosity: it has direct and quantifiable consequences for the analytical sensitivity of any rRNA-targeted assay.

The copy number advantage is very significant. A typical human somatic cell carries approximately 300 to 400 chromosomal rDNA copies distributed across five acrocentric chromosome pairs, with a steady cytoplasmic rRNA pool that reflects constitutive ribosomal biogenesis. An *E. histolytica* trophozoite maintains approximately 200 episomes, each carrying two rDNA copies [20,21]. Unlike chromosomal rDNA, however, these are freely replicating, segregated independently of the cell cycle, and maintained at copy numbers that respond to transcriptional demand. Very importantly, all copies are actively transcribed at the level that is expected of an organism with high biosynthetic requirements during invasive tissue destruction. Such rRNA abundance provides a starting template pool that makes NASBA amplification analytically feasible even from small numbers of organisms or from degraded samples.

Such properties are of particular importance in the ALA context, where liver aspirate is a heterogeneous matrix in which viable trophozoites are concentrated at the margins of the abscess and may be sparse or absent in the central necrotic material. Therefore, the number of intact, RNA-containing parasites in any given aliquot of aspirate may be small. Conventional PCR targeting single-copy genomic sequences would face a sensitivity ceiling under such conditions, while rRNA-targeted NASBA benefits from the multiplicative effect of episomal copy number.

There is yet another dimension to this sensitivity advantage that is specific to RNA targeting. Ribosomal RNA is among the most abundant and metabolically stable RNA species in any actively growing cell. Unlike mRNA, which is subject to rapid turnover and degradation, rRNA is constitutively present in large quantities throughout the cell cycle and retains molecular integrity under conditions that would degrade most other RNA species. In the specific context of sample handling (where a time window exists between sample collection and assay setup) this stability provides a practical margin that is not available to assays targeting less abundant or more labile transcripts.

Taken all together, the episomal architecture of *E. histolytica* rDNA, the high transcriptional activity of all episomal copies, and the inherent stability of the rRNA product converge to provide a natural sensitivity amplification system that operates before the NASBA reaction even begins. The assay exploits a biological property of the pathogen itself to achieve sensitivity that would be difficult to replicate through any amount of additional *in vitro* amplification strategies.

### 5.4. NASBA primer design for AT-rich parasitic targets

The genome of *Entamoeba histolytica* is exceptionally AT-rich, with an overall base composition of approximately 75% adenine and thymine [24]. This is one of the most extreme nucleotide biases recorded among eukaryotic pathogens. This compositional skew propagates into the transcriptome and has direct consequences for the design of oligonucleotide primers targeting *E. histolytica* sequences: primers must anneal within regions where the thermodynamic landscape is dominated by weak AT base pairs, where secondary structure formation in both primer and template is common and the risk of nonspecific annealing to GC-poor off-target sequences is high.

For PCR-based assays operating with high temperature annealing steps, AT richness is managed through extension or primer length, selective enrichment for GC-containing positions within the binding site, and careful optimisation of magnesium concentration and annealing temperature. NASBA presents a fundamentally different thermodynamic context: its cyclic amplification phase operates isothermally at 41°C and primers must achieve stable, productive annealing at this single fixed temperature throughout the entirety of the reaction. There is no freedom to raise the annealing temperature to improve specificity nor to lower it to accommodate weakly binding primers. The primer pair must be designed with melting temperatures sufficiently higher than 41°C to guarantee stable annealing throughout the reaction while remaining close enough to each other in *T*^m^ to ensure balanced competition for their respective templates during the cyclic phase.

In this work, the gene-specific portions of NP1 and NP2 achieve this balance with considerable precision. The calculated melting temperatures (53.81°C and 53.25°C, respectively) place both primers approximately 12°C above the NASBA reaction temperature, therefore providing a robust annealing margin that accommodates the reduced ionic strength and enzyme-mediated structural dynamics of the NASBA reaction without compromising specificity. Likewise, the Δ*T*^m^ of 0.56°C between them is particularly noteworthy: in an AT-rich target context where small sequence changes can produce disproportionate *T*^m^ shifts, achieving such close thermal matching across two independently designed primers targeting different strands of the sample amplicon reflects the identification of an unusually favourable binding site within the SSU rRNA sequence.

The inclusion of 15% v/v dimethylsulphoxide in the reaction buffer addresses a secondary consequence of AT richness: the propensity of AU-rich single-stranded RNA to adopt stable intramolecular secondary structures (hairpins, bulges, etc.) that compete with primer annealing by sequestering the target sequence in folded conformations. DMSO acts as a helix-destabilising agent, lowering the effective melting temperature of secondary structures and increasing the proportion of target RNA available in an accessible single-stranded conformation at 41°C. Its use at 15% is standard in NASBA protocols for structured RNA targets and complements the 65°C denaturation step, which provides an initial unfolding of the RNA template that the lower reaction temperature alone cannot sustain.

Lastly, the T7 promoter prefix appendd to NP1 introduces a further design consideration. The 25-nt T7 sequence (5’-AATTCTAATACGACTCACTATAGGG-3’) does not anneal to the target RNA; it folds back on the newly synthesised cDNA strand during the initiation phase to form the double-stranded promoter recognised by the T7 RNA polymerase. This non-annealing tail does not contribute to the *T*^m^ of NP1 and must be excluded from thermodynamic calculations. The net result is a primer pair whose annealing behaviour in the NASBA reaction is governed entirely by the gene-specific portions, both of which are thermally balanced despite the AT-rich sequence context in which they operate.

### 5.5. Limitations

The findings reported here ought to be interpreted within the constraints of a single case study and several limitations of both the clinical report and the assay itself require explicit acknowledgment.

The most fundamental limitation is the study design. A case report cannot provide estimates of diagnostic sensitivity and specificity. The NASBA assay described here produced a single positive result in a single patient and no conclusion about the performance of the assay across a population of patients can be drawn from this observation alone. Formal analytical validation, including determination of the limit of detection, assessment of reproducibility across independent enzyme preparations, and specificity testing against clinical isolates and extracted RNA of *E. dispar* and other *Entamoeba* species, remains to be performed before the assay can be proposed for routine diagnostic use.

The specificity of the primer pair against non-*E. histolytica Entamoeba* species was confirmed by *in silico* BLAST analysis but no wet laboratory confirmation testing against *E. dispar, E. moshkovskii*, and other clinically relevant species was carried out. While the BLAST result provides strong computational evidence for sequence-level exclusion, empirical confirmation against extracted RNA from characterised isolates is the appropriate standard and was not carried out in the present study.

The in-house nature of the enzyme mixture introduces a further limitation. Unlike commercially validated NASBA kits with standardised enzyme activity specifications, the AMV-RT, RNase H, and T7 RNA polymerase used here were sourced and assembled in house. The batch-to-batch consistency of in-house enzyme preparations is not guaranteed without systematic quality control procedures and the reaction conditions described in Section 3.4 may require re-optimisation if the enzyme sources change. This limits direct transferability of the protocol to other laboratories without independent validation.

The gel electrophoresis readout, while accessible and informative in the proof-of-concept context, carries limitations that preclude point-of-care deployment in its current form. Post-amplification handling of open tubes for gel loading introduces a carryover contamination risk that is particularly relevant in this context were the RNA amplicon is itself a highly efficient template for re-amplification. Any translation of this assay to field or primary care settings would require conversion to a closed-tube detection format such as lateral flow assay or molecular beacon, as discussed in Section 3.4.

Finally, the internal control design, while robust against inhibition, has an inherent interpretive limitation. The *Saccharina japonica* rbcL control monitors amplification and enzyme integrity; a visible control band confirms that the reaction proceeded. However, it cannot distinguish between a true negative result (genuine absence of *E. histolytica* RNA in the sample) and a false negative caused by RNA degradation in the aspirate between the time of collection and the time of extraction. Pre-analytical RNA integrity is a known vulnerability of RNA-targeted molecular assays in clinical practice and the time elapsed between aspiration and processing was not formally recorded in this case. Inclusion of a specimen adequacy control (an endogenous RNA target present in all clinical material regardless of the pathogen) would strengthen the interpretive framework in future implementations.

### 5.6. Implications for point-of-care diagnostics in NTD-endemic settings

The ASSURED criteria – Affordable, Sensitive, Specific, User-friendly, Rapid/robust, Equipment-free, and Delivered – define the aspirational standard for point-of-care diagnostics in resource-limited settings, as articulated by the World Health Organisation [25]. No single technology simultaneously satisfies all criteria for every target, but the platform described here, particularly when coupled with lateral flow assay readout, aligns with this framework to a degree that few molecular diagnostic approaches for intestinal and hepatic parasitoses can match.

The isothermal operating temperature of 41°C is achievable with a simple dry block heater, a water bath, or even a controlled temperature incubator of the kind found in primary care laboratories. Unlike PCR, NASBA requires no thermal cycling, no ramp rate calibration, and no voltage-stable power supply capable of sustaining rapid temperature transitions. The reaction chemistry is complete in 90 minutes from sample addition to readout, which is a turnaround that is compatible with same-day clinical decision-making in an outpatient or emergency setting. These properties address the infrastructure barrier that currently confines species-level molecular confirmation of *Entamoeba* infection to reference or university laboratories, removing it from the care pathway of the majority of patients who present with ALA in endemic regions.

The case for LFA-based readout in this context is compelling. The conversion from gel electrophoresis to a lateral flow strip format requires no changes to the primer sequences, reaction chemistry or amplification protocol. It is a detection-only modification accomplished by introducing a 5’ biotin label on NP2 and deploying a nitrocellulose strip with an immobilised capture probe complementary to an internal region of the sense strand amplicon. The result is a visual readout that requires no instrumentation, no electricity at the detection step, and no trained laboratory technician to interpret it. In the context of a district hospital in a low-income country or a primary care unit in a rural endemic zone, this difference between electrophoresis and LFA is the difference between a test that can be performed and one that cannot.

Brazil presents a specific and immediate context for this argument. Amoebiasis remains endemic across the northern, northeastern, and centre-west regions, with prevalence of *Entamoeba spp*. reaching 50% in some states and the pathogenic *E. histolytica* accounting for a meaningful fraction of molecularly confirmed infections [8]. Within its Unified Health System (SUS), the pressure to rationalise antiamoebic prescribing is real. Metronidazole is freely dispensed, its use in *E. dispar* carriers is common, but the infrastructure for species-level confirmation is absent at most primary care levels. An isothermal RNA amplification assay with LFA readout, validated against the Brazilian *E. histolytica* strain background and deployable with minimal cold chain requirements would fill a diagnostic gap that existing tools have not closed.

The broader NTD context is equally relevant. Amoebiasis remains absent from the WHO official NTD list, a classification gap that has attracted sustained criticism given its global burden of morbidity and mortality [2]. The availability of a simple, deployable molecular test for *E. histolytica* species confirmation would strengthen the case for inclusion by enabling accurate epidemiological data collection in endemic settings where the distinction between species has historically been invisible to surveillance systems relying on microscopy. Diagnostics and disease burden estimates are mutually reinforcing: better tools generate better data and better data generates the advocacy that drives resource allocation. The assay described here, in its current proof-of-concept form, represent a step towards that translational objective.

## 6. Conclusions

We describe the design, molecular rationale, and clinical application of a NASBA assay targeting the 40S small subunit ribosomal RNA of *Entamoeba histolytica*, deployed for urgent species-level diagnosis in a patient with amoebic liver abscess with secondary cholestatic hepatic biochemical derangement. The assay produced a positive result that supported the continuation of targeted antiamoebic therapy, resolving a diagnostic dilemma in which the competing risks of empirical treatment and withholding treatment were both clinically significant.

The choice of the 40S SSU rRNA as target exploits two properties unique to *E. histolytica* among eukaryotic pathogens: the extrachromosomal episomal architecture of its ribosomal DNA, which generates an endogenous pre-analytical amplification of template abundance, and the isothermal RNA-selective mechanism of NASBA, which provides mechanistic exclusion of double-stranded DNA signal independently of primer design. These two properties, combined with sequence-level species discrimination confirmed by *in silico* analysis against the *Entamoeba* genus, lead to a unique testing situation. In it, two orthogonal and simultaneously operating layers confer enhanced specificity that is particularly suited to the complex matrix of liver abscess aspirate, thus making it not merely advantageous but clinically necessary.

In its current gel electrophoresis format, the assay constitutes a proof of concept. Conversion to lateral flow assay or molecular beacon readout would remove the remaining infrastructure hurdles and position NASBA-based *E. histolytica* detection as a genuinely deployable point-of-care tool for endemic settings. Formal analytical validation, including the determination of limit of detection, cross-reactivity testing against characterised *Entamoeba* isolates, and multicentre clinical evaluation against a reference standard, represents the necessary step towards that objective.

## Data Availability

As a case report, all the relevant data is included in the manuscript itself.

## References

[1] World Health Organization. Amoebiasis. Wkly Epidemiol Rec. 1997;72(14):97–99.

[2] Stanley SL. Amoebiasis. Lancet. 2003 Mar 22;361(9362):1025–1034.

[3] Fotedar R, Stark D, Beebe N, Marriott D, Ellis J, Harkness J. Laboratory diagnostic techniques for Entamoeba species. Clin Microbiol Rev. 2007 Jul;20(3):511–532.

[4] Haque R, Huston CD, Hughes M, Houpt E, Petri Jr WA. Amebiasis. New Engl J Med. 2003 Apr 17;348(16):1565–1573.

[5] Pritt BS, Clark CG. Amebiasis. In Mayo Clin Proc 2008 Oct 1; 83(10): 1154–1160.

[6] Blessmann J, Van Linh P, Nu PA, Thi HD, Muller-Myhsok B, Buss H, Tannich E. Epidemiology of amebiasis in a region of high incidence of amebic liver abscess in central Vietnam. Am J Trop Med Hyg. 2002 May 1;66(5):578–583.

[7] Diamond LS, Clark CG. A Redescription of Entamoeba Histolytica Schaudinn, 1903 (Emended Walker, 1911) Separating It From Entamoeba dispar Brumpt, 1925 1. J Eukaryot Microbiol. 1993 May;40(3):340–344.

[8] dos Santos Zanetti A, Malheiros AF, de Matos TA, Dos Santos C, Battaglini PF, Moreira LM, Lemos LM, Castrillon SK, Cortela DD, Ignotti E, Espinosa OA. Diversity, geographical distribution, and prevalence of Entamoeba spp. in Brazil: a systematic review and meta-analysis. Parasite. 2021 Mar 30;28:17.

[9] Brumpt E. Étude sommaire de l’Entamoeba dispar n. sp. Amibe à kystes quadrinucléés. Bull Acad Méd. 1925:94:943–945.

[10] Al-Hussaniy HA. Exploring the safety and adverse effects of metronidazole. Iraqi J Pharm. 2024 Jan 30;1(1):1–2.

[11] Fotedar R, Stark D, Beebe N, Marriott D, Ellis J, Harkness J. Laboratory diagnostic techniques for Entamoeba species. Clin Microbiol Rev. 2007 Jul;20(3):511–532.

[12] Knobloch J, Mannweiler E. Development and persistence of antibodies to Entamoeba histolytica in patients with amebic liver abscess. Analysis of 216 cases. Am J Trop Med Hyg. 1983:32(4):727–732.

[13] Haque R, Mollah NU, Ali IK, Alam K, Eubanks A, Lyerly D, Petri Jr WA. Diagnosis of amebic liver abscess and intestinal infection with the TechLab Entamoeba histolytica II antigen detection and antibody tests. J Clin Microbiol. 2000 Sep 1;38(9):3235–3239.

[14] Roy S, Kabir M, Mondal D, Ali IK, Petri Jr WA, Haque R. Real-time-PCR assay for diagnosis of Entamoeba histolytica infection. J Clin Microbiol. 2005 May; 43(5):2168–2172.

[15] Srivastava P, Prasad D. Isothermal nucleic acid amplification and its uses in modern diagnostic technologies. 3 Biotech. 2023 Jun;13(6):200.

[16] Compton J. Nucleic acid sequence-based amplification. Nature. 1991;350(6313):91–92.

[17] Deiman B, van Aarle P, Sillekens P. Characteristics and applications of nucleic acid sequence-based amplification (NASBA). Mol Biotech. 2002 Feb;20(2):163–179.

[18] Mugasa CM, Laurent T, Schoone GJ, Basiye FL, Saad AA, El Safi S, Kager PA, Schallig HD. Simplified molecular detection of Leishmania parasites in various clinical samples from patients with leishmaniasis. Parasites & Vectors. 2010 Mar 2;3(1):13.

[19] Deborggraeve S, Büscher P. Molecular diagnostics for sleeping sickness: what is the benefit for the patient?. Lancet Infect Dis. 2010 Jun 1;10(6):433–439.

[20] Huber M, Koller B, Gitler C, Mirelman D, Revel M, Rozenblatt S, Garfinkel L. Entamoeba histolytica ribosomal RNA genes are carried on palindromic circular DNA molecules. Mol Biochem Parasitol. 1989 Jan 15;32(2-3):285–296.

[21] Bhattacharya S, Bhattacharya A, Diamond LS, Soldo AT. Circular DNA of Entamoeba histolytica encodes ribosomal RNA. J Protozool. 1989 Sep;36(5):455–458.

[22] Hamzah Z, Petmitr S, Mungthin M, Leelayoova S, Chavalitshewinkoon-Petmitr P. Differential detection of Entamoeba histolytica, Entamoeba dispar, and Entamoeba moshkovskii by a single-round PCR assay. J Clin Microbiol. 2006 Sep;44(9):3196–3200.

[23] Tyagi S, Kramer FR. Molecular beacons: probes that fluoresce upon hybridization. Nature Biotechnol. 1996 Mar 1;14(3):303–308.

[24] Loftus B, Anderson I, Davies R, Alsmark UC, Samuelson J, Amedeo P, Roncaglia P, Berriman M, Hirt RP, Mann BJ, Nozaki T. The genome of the protist parasite Entamoeba histolytica. Nature. 2005 Feb 24;433(7028):865–868.

[25] Peeling RW, Holmes KK, Mabey D, Ronald A. Rapid tests for sexually transmitted infections (STIs): the way forward. Sexually Transmitted Infections. 2006 Dec 1;82(suppl 5):v1–6.

